# Post-stroke executive function in relation to white matter damage on clinically acquired CT brain imaging

**DOI:** 10.1101/2021.11.12.21266247

**Authors:** Georgina Hobden, Margaret Moore, Grant Mair, Sarah Pendlebury, Nele Demeyere

## Abstract

**Importance:** Executive function impairments are highly prevalent post-stroke and are a feature of vascular dementia. Computed tomography (CT) neuroimaging is routinely acquired in clinical settings. Therefore, determining the prognostic utility of CT-derived markers for post-stroke cognitive outcomes is of key clinical and academic interest.

**Objective:** To determine whether post-stroke executive function is associated with stroke-related white matter damage and/or white matter hypoattenuations of presumed vascular origin (WMHs) on routine CT brain scans.

**Design:** Retrospective cross-sectional analysis of data collected within the Oxford Cognitive Screening (OCS) programme (2016-2020).

**Setting:** Patients were recruited at Oxford University Hospital’s acute stroke unit. Follow-up cognitive assessment was conducted six-months post-stroke at patients’ homes.

**Participants:** OCS programme recruited a consecutive patient sample with a confirmed diagnosis of stroke, who were minimum 18 years old, able to remain alert for 20 minutes, and able to provide informed consent. This study included all patients who completed six-month follow-up assessment with the Oxford Cognitive Screen-Plus (OCS-Plus) and had a usable acute CT scan with a visible stroke lesion.

**Main Outcome and Measures:** Association between post-stroke executive function and both stroke-specific white matter damage and WMHs on routine CT. Executive function was evaluated using the OCS-Plus Rule Finding task. Stroke lesions were manually delineated on CT, and stroke-related white matter damage was quantified then dichotomised using the HCP-842 atlas. WMHs were visually rated using the Age-Related White Matter Changes scale and dichotomised as present or absent.

**Results:** Among 87 stroke patients (mean/SD age = 73.60/11.75; 41 female; 61 ischaemic stroke), multivariable linear regression analyses demonstrated that poorer executive function six-months post-stroke was associated with both stroke damage to the medial lemniscus (*B*= - 8.86, *p*< .001, 95% CI [-13.29 -4.43]) and the presence of WMHs (*B*= -5.42, *p*= .005, 95% CI [-9.12 -1.72]), after adjusting for covariates including age and education.

**Conclusions and Relevance:** Poorer post-stroke executive functioning was associated with both localised patterns of stroke-specific white matter damage and white matter degeneration. Our results confirm the necessary role of white matter integrity for executive functioning post-stroke and highlight the prognostic utility of CT-derived neuroimaging markers for long-term post-stroke cognitive outcomes.

## INTRODUCTION

Stroke is the third most important cause of disability burden worldwide.^1,2^ Whilst stroke mortality rates have decreased due to improved acute care,^3^ there is a higher prevalence of chronic stroke survivors^4^ and prevalence of post-stroke cognitive impairment is high.^5^ In particular, executive dysfunction is estimated to affect between 19% and 75% of chronic stroke survivors.^6,7,8^ Post-stroke executive function impairment has been associated with reduced quality of life,^9^ disability in activities of daily living,^10^ and increased mortality.^11^

Previous studies using magnetic resonance imaging (MRI) have linked post-stroke executive dysfunction to focal lesions affecting white matter pathways^12^ and white matter hyperintensities of presumed vascular origin (WMHs),^13,14^ which affect 64-86% of stroke patients.^15,16^ While these published studies provide valuable insight into the neural mechanisms underpinning post-stroke executive dysfunction, they yield comparatively ungeneralisable results due to their reliance on MRI, which is not always suitable for a significant, non-random portion of the stroke population (e.g., patients with implanted paramagnetic devices).^17^ Furthermore, previous studies have investigated the role of stroke-specific white matter damage or WMHs in isolation but have not studied how these conditions interact in the same patient sample.

The present investigation used CT brain scans collected as part of routine clinical care to investigate the association between post-stroke executive function and both pre-and post-stroke white matter damage. Using routinely acquired CT imaging allowed us to investigate these associations in a relatively representative patient sample. Furthermore, as CT is the most commonly used imaging modality in acute stroke settings, it is critical to determine its prognostic utility for post-stroke outcomes. In line with previous MRI studies, we hypothesised that post-stroke executive dysfunction would be predicted by CT-derived measures of both stroke-specific white matter damage and WMHs.

## METHODS

### Standard Protocol Approvals, Registrations, and Patient Consents

This project is a retrospective analysis of data collected within the Oxford Cognitive Screening (OCS) programme,^18^ which recruited a consecutive sample of stroke survivors during acute hospitalisation and conducted follow up neuropsychological assessments at six-months. The OCS programme received regional ethics approval (OCS-Tablet and OCS-Recovery studies, NHS RECs 14/LO/0648 and 18/SC/0550). All patients provided written or witnessed informed consent at recruitment and follow-up.

### Participants

The OCS programme recruited a consecutive sample of stroke patients at the Oxford University Hospital’s acute stroke unit and included all patients with a confirmed diagnosis of acute stroke, who were at least 18 years old, were able to remain alert for 20 minutes, and were able to provide informed consent.

The present investigation included all patients recruited for the OCS programme that had both completed the follow-up assessment with the more detailed Oxford Cognitive Screen-Plus (OCS-Plus)^19^ six-months after stroke (completed with participants between November 2016-March 2020), and had a usable CT scan from the acute stage post-stroke (i.e., within 0-14 days post-stroke) with a visible stroke lesion (*n* = 122). Patients were excluded if their CT scan showed evidence of additional non-stroke pathology (e.g., brain tumour: *n* = 3), or multiple temporally distinct strokes (*n* = 20), or if the Rule Finding task was not completed (*n* = 12). Demographic and clinical details of the resultant sample of 87 stroke survivors as reported by relevant medical records are presented in Table 1.

**Table 1.** A breakdown of patient demographic and clinical information as recorded by medical records.

| **Characteristics** | **Statistics** |
| --- | --- |
| Age, *M (SD)* | 73.60 (11.75) |
| Education, *M (SD)* | 12.74 (3.67) |
| Sex, *N (%)* | Female: 41 (47.13); Male: 46 (52.87) |
| Handedness, *N (%)* | Right: 58 (66.67); Left: 4 (4.60); Unknown: 25 (28.74) |
| Stroke type, *N (%)* | Ischaemic: 61 (70.11); Haemorrhagic: 26 (29.89) |
| Stroke lateralisation, *N (%)* | Unilateral right hemisphere: 43 (49.43); Unilateral left hemisphere: 41 (47.13); Bilateral*: 3 (3.45) |
| Stroke location, *N (%)* | Anterior cerebral artery: 3 (3.45); Middle cerebral artery: 61 (70.11); Posterior cerebral artery: 8 (9.20); Basilar artery: 7 (8.05); Intraventricular: 3 (3.45); Multiple territories: 5 (5.75) |
| Stroke volume (cm<sup>3</sup>), *MED (IQR)* | 10.82 (44.89) |
| Days between stroke and scan, *M (SD)* | 0.94 (2.31) |

\*Bilateral stroke was defined as a single stroke crossing the midline.
*SD* = standard deviation.
*MED* = median.
*IQR* = interquartile range.

### Cognitive data

Executive function was assessed six-months after stroke using the OCS-Plus, a tablet-based cognitive screen designed to provide fine-grained measures of memory and executive function.^19^ This tool has been validated versus standardised pen-and-paper neuropsychological assessments in a large normative aging sample (*n* = 320).^19^ The OCS-Plus contains 10 subtasks and takes on average 24 minutes to complete.^19^ OCS-Plus subtasks are designed to tap cognitive subdomains with minimal interference from impairments within separate cognitive domains (e.g., language, memory).^20^

We evaluated data from the OCS-Plus Rule Finding task, which was designed to assess complex executive function.^19^ The Rule Finding task is analogous to the Brixton Spatial Anticipation test,^21^ but it can be administered more quickly and places lower demands on working memory. Patients are shown three columns of squares and triangles which regularly vary in luminosity (Supplementary Figure 1). A red dot is placed within a shape, then moves throughout the array according to a specific but changing spatial rule. Patients are instructed to learn the rule guiding the dot’s movement to predict where the dot will appear next. The Rule Finding task thus taps executive function by requiring patients to update their working memory representation of the spatial pattern, to apply new spatial rules to make predictions once a new rule has been learned, and to inhibit obsolete rules. Performance accuracy was scored based on the number of correct location predictions made (range = 0-43).^19^

### Lesion analyses

Acute whole-brain non-enhanced CT scans (slice thickness 5mm) were collected for each patient. Stroke lesions were manually delineated on native space scans by two experienced neuroimaging researchers (GH and MM) using the MRIcron software package (McCausland Center for Brain Imaging, Columbia, SC, USA) and following a standardised processing procedure.^22^ Researchers were blinded to behavioural and clinical data. The resultant lesion masks were evaluated for accuracy by a consultant neuroradiologist with over 13 years’ experience (GM), who advised on minor adjustments to the lesion masks. Following GM’s evaluation, lesion masks were smoothed at 5 mm full width at half maximum in the z-direction and binarized using a threshold of 0.5. Scans and lesion masks were reoriented to the anterior commissure and warped into 1×1×1mm stereotaxic space using the Statistical Parametric Mapping 12 and Clinical Toolbox.^23^ Normalised scans and lesion masks were visually inspected for quality.

Binarised lesion masks were used to calculate stroke volume and white matter disconnection severity statistics. Post-stroke white matter damage was quantified in terms of white matter tract disconnection severities, which were estimated using the Lesion Quantification Toolkit.^24^ This toolkit estimates tract-level disconnection for 70 canonical white matter tracts, as defined by the HCP-842 streamline tractography atlas,^25^ with the corpus callosum divided into five segments based on Freesurfer corpus callosum segmentation. The toolkit works on CT by embedding the lesion into the HCP-842 as a region of interest (ROI) and iteratively loading the streamline trajectories for each of the tracts. The files are filtered to retain only those streamlines that intersect the volume occupied by the lesion.

For each tract, we calculated the percentage disconnection severity by converting the number of disconnected streamlines into a percentage of the total number of streamlines assigned to that tract. Given that anatomical homologues of some white matter ROIs exist within both hemispheres, we calculated separate metrics for each of these homologues, then averaged across hemispheres to produce a single measure per patient for each ROI. Each ROI was then binarized as disconnected (at least 10% disconnection) or not disconnected (less than 10% disconnection).^24^ In addition to these individual tract-level measures, we calculated a measure of white matter disconnection to reflect the extent of stroke-specific disconnection across the brain by summing the number of disconnected ROIs for each patient (range = 0-40).

### WMH ratings

WMHs were evaluated on the clinical CT scans using the Age-Related White Matter Changes visual rating scale (ARWMC scale).^26^ Visual ratings were assigned by an experienced neuroimaging researcher (GH), who was blinded to behavioural and clinical data. WMHs were assessed in five regions within each hemisphere (10 regions total): frontal, parieto-occipital, temporal, infratentorial, and basal ganglia. Each region was assigned a score of 0-3 according to the criteria in Supplementary Table 1. In cases where a brain region was severely affected by stroke damage so that WMHs could not be evaluated, the affected region was assigned the same score as the anatomically homologous region in the opposite hemisphere. A total score reflecting the severity of WMHs across the brain was calculated by summing the assigned scores across all ten regions (range = 0-30). This total score was used to categorise WMHs as absent (0), mild (1-5), moderate (6-10), or severe (>10).^27^ WMHs were then dichotomised as present or absent for subsequent statistical analyses due to statistical power concerns. Consultant neuroradiologist (GM) independently evaluated WMHs to increase robustness of the evaluations. Cases where GH’s and GM’s evaluations differed were revisited and discussed to reach a final consensus prior to statistical analysis.

### Statistical analysis

All statistical analyses were performed using the computer software R-Studio 4.0.2. Behavioural and clinical data were cleaned before analysis. Mean substitution was employed to impute missing values for patient age (*n* = 7) and education (*n* = 30).

Multivariable linear regression analyses evaluated post-stroke executive function in relation to: (1) stroke-specific damage to white matter ROIs across the brain; (2) stroke damage to individual white matter ROIs; (3) non-stroke white matter damage (WMHs). Finally, we evaluated the relative contributions of both stroke-specific white matter damage and WMHs to post-stroke executive function by conducting a multivariable linear regression analysis, which included as predictor variables any significant predictors of executive function identified in the original analyses (i.e., analyses 1, 2, 3). For all multivariable linear regression analyses, the OCS-Plus Rule Finding executive function score was used as the outcome measure. All regression analyses included age, sex, years of education, stroke type (ischaemic, haemorrhagic), stroke lateralisation (left, right, bilateral), stroke territory (anterior cerebral artery, middle cerebral artery, posterior cerebral artery, vertebrobasilar, intraventricular, multiple territories) and stroke volume as covariates of no interest.

Residual plots were visually inspected to test the assumptions of linearity, normality of residuals, and homogeneity of residuals variance for all multivariable linear regression analyses. Where appropriate, the Bonferroni correction was applied to correct for inflated false positive rates due to multiple comparisons.

### Data Availability

Data used in the analyses of the present study have been made openly available on the Open Science Framework (https://osf.io/r4h3m/). The CT brain scans cannot be shared openly due to ethical research governance restrictions on the clinical imaging data.

## RESULTS

First, basic descriptive analyses were conducted to investigate data from the behavioural, lesion, and visual rating measures employed in this study. The mean score for the OCS-Plus Rule Finding task was 18.29 (*SD* = 8.29, range = 2-37). Figure 1 presents a lesion overlay for the sample. Table 2 shows the prevalence and extent of ROI disconnection for the sample. Overall, there was a good level of lesion coverage across ROIs considered in the present investigation. The mean number of ROIs disconnected by at least 10% was 9.79 (*SD* = 8.97, range = 0-34). WMHs were categorised as absent, mild, moderate, or severe in 32, 31, 15, and 9 patients, respectively. Due to concerns about statistical power given these relatively small subgroups, WMHs were binarized as absent (*n* = 32) or present (*n* = 55) for subsequent statistical analyses.

**Figure 1.**
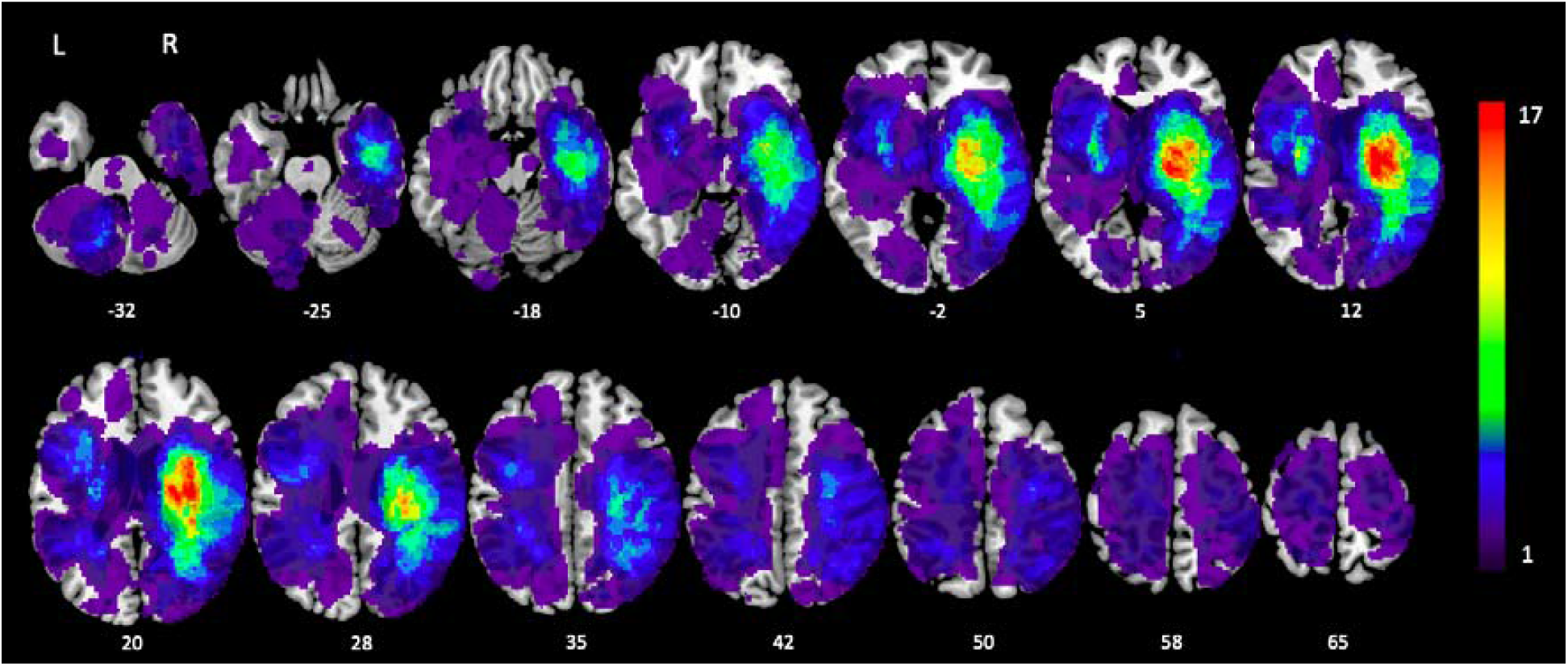
A lesion overlay map of the study sample (*n* = 87) presented in neurological convention. Colour indicates the number of patients with lesions in each area. MNI coordinates of each transverse section (z-axis) are provided.

**Table 2.** Descriptive statistics for each white matter region of interest from the HCP-842 analysed in the present investigation.

| Region of interest | Patients with any ROI disconnection, *N* (%) | Patients with >10% ROI disconnection, *N* (%) | Average percentage disconnection, *M* (*SD*) |
| --- | --- | --- | --- |
| Anterior commissure | 41 (47.13) | 31 (35.63) | 25.66 (38.81) |
| Arcuate fasciculus | 45 (51.72) | 37 (42.53) | 17.62 (22.09) |
| Acoustic radiation | 38 (43.68) | 30 (34.48) | 14.90 (20.97) |
| Cerebellum | 14 (16.09) | 5 (5.75) | 1.49 (4.75) |
| Corpus callosum (anterior) | 34 (39.08) | 14 (16.09) | 6.03 (13.24) |
| Corpus callosum (mid-anterior) | 41 (47.13) | 24 (27.59) | 11.83 (24.49) |
| Corpus callosum (central) | 31 (35.63) | 22 (25.29) | 14.56 (29.95) |
| Corpus callosum (mid-posterior) | 44 (50.57) | 21 (24.14) | 16.36 (32.81) |
| Corpus callosum (posterior) | 61 (70.11) | 28 (32.18) | 21.22 (33.48) |
| Corticospinal tract | 57 (65.52) | 40 (45.98) | 18.80 (21.66) |
| Corticostriatal tract | 72 (82.76) | 33 (37.93) | 11.01 (13.41) |
| Central tegmental tract | 8 (9.20) | 7 (8.05) | 5.30 (18.88) |
| Corticothalamic pathway | 77 (88.51) | 40 (45.98) | 13.79 (16.30) |
| Cingulum | 28 (32.18) | 15 (17.24) | 5.38 (12.41) |
| Dorsal longitudinal fasciculus | 7 (8.05) | 7 (8.05) | 6.18 (22.06) |
| Extreme capsule | 47 (54.02) | 40 (45.98) | 19.68 (22.01) |
| Frontal aslant tract | 30 (34.48) | 25 (28.74) | 7.14 (23.15) |
| Frontopontine tract | 52 (59.77) | 37 (42.53) | 16.81 (20.48) |
| Fornix | 17 (19.54) | 16 (18.39) | 9.02 (22.41) |
| Inferior cerebellar peduncle | 8 (9.20) | 8 (9.20) | 3.18 (11.19) |
| Inferior fronto-occipital fasciculus | 53 (60.92) | 37 (42.53) | 17.80 (22.67) |
| Inferior longitudinal fasciculus | 33 (37.93) | 22 (25.29) | 10.03 (18.49) |
| Lateral lemniscus | 4 (4.60) | 4 (4.60) | 1.86 (9.14) |
| Middle cerebellar peduncle | 19 (21.84) | 5 (5.75) | 1.65 (6.96) |
| Medial longitudinal fasciculus | 6 (6.90) | 5 (5.75) | 4.16 (17.64) |
| Medial lemniscus | 22 (25.29) | 20 (22.99) | 9.58 (18.16) |
| Middle longitudinal fasciculus | 35 (40.23) | 29 (33.33) | 15.49 (22.17) |
| Occipitopontine tract | 43 (49.43) | 32 (36.78) | 16.89 (22.45) |
| Optic radiation | 36 (41.38) | 25 (28.74) | 12.41 (21.18) |
| Posterior commissure | 16 (18.39) | 7 (8.05) | 7.09 (25.09) |
| Parietopontine tract | 51 (58.62) | 38 (43.68) | 17.60 (21.28) |
| Reticulospinal tract | 6 (6.90) | 6 (6.90) | 2.61 (10.38) |
| Superior cerebellar peduncle | 27 (31.03) | 14 (16.09) | 8.21 (18.62) |
| Superior longitudinal fasciculus | 37 (42.53) | 23 (26.44) | 9.72 (16.30) |
| Spinothalamic tract | 25 (28.74) | 21 (24.14) | 9.07 (17.08) |
| Temporopontine tract | 36 (41.38) | 28 (32.18) | 15.15 (21.73) |
| Uncinate fasciculus | 26 (29.89) | 16 (18.39) | 7.75 (16.58) |
| U-fibres | 61 (70.11) | 23 (26.44) | 7.68 (10.95) |
| Vermis | 9 (10.34) | 7 (8.05) | 6.26 (22.60) |
| Vertical occipital fasciculus | 16 (18.39) | 10 (11.49) | 4.40 (12.16) |

Next, we conducted a multivariable linear regression analysis to investigate the relationship between post-stroke executive function and stroke-specific white matter damage using the overall white matter disconnection score (number of ROIs disconnected by at least 10%). The executive function score was not significantly predicted by the overall stroke-specific white matter disconnection measure, although age and stroke volume were significant negative predictors of executive function. See Supplementary Table 2 for full model statistics.

Next, we conducted multivariable linear regressions to assess the relationship between post-stroke executive function and individual ROI-level tract disconnection severity. Post-stroke executive function was associated with damage to the posterior corpus callosum, inferior longitudinal fasciculus, middle cerebellar peduncle, medial lemniscus, occipitopontine tract, parietopontine tract, spinothalamic tract, temporopontine tract, and vermis before applying the Bonferonni correction for multiple comparisons. However, after applying the Bonferroni correction for multiple comparisons, only the association between post-stroke executive function and damage to the medial lemniscus remained. (Figure 2a). Age and stroke volume were significant negative predictors of post-stroke executive function in all analyses. See Supplementary Table 2 for full model statistics.

**Figure 2.**
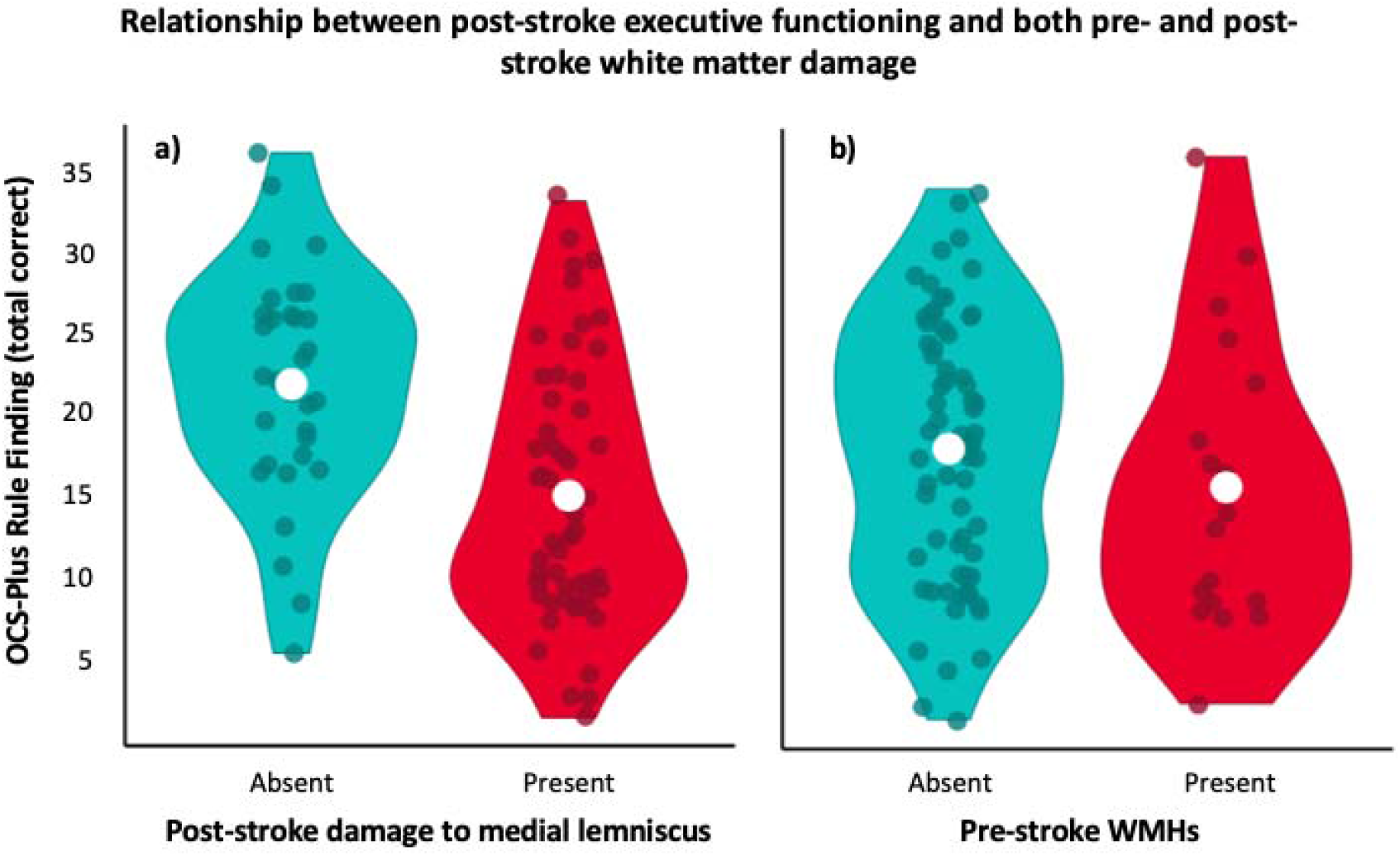
The relationship between post-stroke executive function and (a) stroke damage to the medial lemniscus, and (b) WMHs. Executive function was measured using the OCS-Plus Rule Finding task. Stroke damage to the medial lemniscus and WMHs were assessed on clinical CT brain imaging. Means are plotted (in white) along with individual patient scores (points).

Next, we conducted a multivariable linear regression to investigate the relationship between post-stroke executive function score and the presence of WMHs. Post-stroke executive function was significantly poorer in patients with WMHs, compared to patients without WMHs (*B* = -4.17, *p* = .042, 95% CI [-8.18 -0.15]). (Figure 2b). Additionally, age and stroke volume were significant predictors of lower executive function scores. No other covariates were significant. See Supplementary Table 2 for full model statistics.

Finally, we conducted a multivariable linear regression to investigate the relative contributions of stroke-specific white matter damage and WMHs to post-stroke executive function. This model included WMHs (absent vs present) and medial lemniscus damage (absent vs present) as predictor variables of interest. This model was found to be significant (*F*(18, 68) = 3.86, *R*^*2*^ = .51, *R*^*2*^_*Adjusted*_ = .37, *p* < .001). Post-stroke executive function was associated with both stroke damage to the medial lemniscus (*B* = -8.86, *p* < .001, 95% CI [-13.29 -4.43]) and the presence of WMHs (*B* = -5.42, *p* = .005, 95% CI [-9.12 -1.72]), although there was no significant interaction effect between the presence of stroke damage to the medial lemniscus and the presence of WMHs.

### DISCUSSION

This study investigated the association between post-stroke executive function and both pre- and post-stroke white matter damage in a clinically representative sample of chronic stroke survivors using CT brain imaging collected as part of routine clinical care. We found poorer executive functioning in patients with stroke damage affecting the medial lemniscus and in patients with WMHs. These findings highlight the impact of both stroke damage to specific white matter tracts and pre-stroke small vessel disease on post-stroke cognitive outcomes. Furthermore, our results highlight the prognostic utility of CT-derived neuroimaging measures for investigating cognitive outcomes after stroke.

First, stroke-specific white matter damage to the medial lemniscus was associated with significantly poorer executive function six-months after stroke. The medial lemniscus is a key white matter pathway that connects the cerebellum and thalamus, and is considered a critical part of the cerebello-thalamo-cortical pathway between the prefrontal cortex and cerebellum.^28,29^ Our findings suggest that stroke damage to long-range prefrontal-thalamic-sensorimotor projections may be an important determinant of post-stroke executive dysfunction. Although we did not find significant associations between post-stroke executive function and stroke damage to any other white matter pathways that have previously been implicated in executive functioning (e.g., inferior longitudinal fasciculus)^30^, this may be explained by our conservative approach to multiple comparison correction, lack of statistical power in analyses of some tracts that were rarely damaged, and differences in our patient sample compared to previous studies. Indeed, previous MRI studies that have linked executive functioning to specific white matter tracts have generally included younger participant samples without contraindications to MRI,^31,32,33,34^ in contrast to our study that used a largely unselected and therefore more clinically representative cohort.

The presence of WMHs on routinely collected CT brain scans was also associated with significantly poorer executive function six-months after stroke. This finding is critically important as it suggests that post-stroke executive dysfunction may not be caused exclusively by patterns of stroke-specific damage, but may also be linked to large-scale network integrity issues caused by cerebral small vessel disease. The present investigation adds to previous studies that have reported an association between WMHs and post-stroke cognitive outcomes^5,13,14,35^ by demonstrating that WMHs remain a key predictor of post-stroke executive function, even when stroke-specific white matter damage is taken into account.

Our results highlight the potential of routinely acquired CT brain scans for use in clinical research and clinical practice. Although MRI has traditionally been the favoured modality in clinical research, due to its higher spatial resolution and improved soft tissue contrast compared to CT,^36^ the present study demonstrates that CT detects levels of white matter damage that correlate meaningfully with post-stroke cognitive outcome, even after adjusting for important covariates (e.g., age, education). Future studies should further assess the prognostic utility of CT-derived neuroimaging measures for predicting clinical, cognitive, and functional outcomes in acutely unwell patient populations, such as patients with stroke and delirium.^37,38^ Once the prognostic utility of clinical CT scans has been established more firmly, data from acute CT neuroimaging could be incorporated into clinical risk prediction algorithms for post-stroke outcome, particularly should automated quantitative assessment methods become widely clinically available for CT. Such algorithms would facilitate identification of individual patients at risk of poor cognitive and functional outcomes post-stroke.

Several potential limitations are present within this study. First, our assessment of post-stroke white matter damage and WMHs may have been complicated by the presence of cerebral oedema, a common sequela of stroke. Nevertheless, oedema typically peaks 3-5 days after stroke^39^ and only six CT scans in the present investigation were acquired during or after this period. Second, we combined acute imaging data with chronic behavioural data, which may have weakened statistical lesion-behaviour relationships, as patients who exhibited executive dysfunction at the acute stage may have recovered by the time of chronic assessment. Future investigations should therefore determine whether our results generalise to stroke patients’ executive functioning in the acute stage post-stroke. Third, we used a different measurement approach to assess post-stroke white matter damage and WMHs, as we assessed the former using a tract disconnection measure and the latter using a visual rating approach. This was necessitated by the current lack of openly available quantitative tools to assess WMHs on CT, despite the availability of such tools for MRI,^40^ and by the present study’s sample size, which left us underpowered to investigate any dose-responsive effect of WMHs. Nevertheless, we found significant results for both pre- and post-stroke white matter damage, suggesting that both approaches were sufficiently sensitive in correlation with cognitive data. Finally, while we aimed to include a clinically representative sample by recruiting consecutively from a regional stroke unit, the need for informed consent and 6-month follow-up likely resulted in some selection bias.

Overall, this investigation confirms the association between post-stroke executive function and both stroke-specific white matter damage and WMHs in a clinically representative patient sample using routine clinical CT brain imaging. Our results demonstrate the influence of both stroke damage and cerebral small vessel disease-related white matter damage on post-stroke cognitive outcomes. Furthermore, our results have implications for clinical research and clinical practice, by demonstrating the prognostic utility of CT-derived neuroimaging measures for predicting post-stroke cognitive outcomes.

## Data Availability

All data used are made available online at the Open Science Framework.

https://osf.io/r4h3m/

## ACKNOWLEDGEMENTS

This study was supported by a Priority Programme Grant from the Stroke Association (SA PPA 18/100032). GH is supported by ESRC doctoral training network funding. GM is the Stroke Association Edith Murphy Foundation Senior Clinical Lecturer (SA L-SMP 18\1000). The study was further supported by the National Institute for Health Research (NIHR) the Clinical Research Network and the NIHR Oxford Biomedical Research Centre (BRC) based at Oxford University Hospitals NHS Trust and University of Oxford. The views expressed are those of the author(s) and not necessarily those of the NHS, the NIHR or the Department of Health.

We would like to thank the participants who to part in the Oxford Cognitive Screening study and all research staff who contributed towards data collection. In particular, we acknowledge the contributions to data collection and curation for the OCS data made by Ms Ellie Slavkova, Ms Grace Chiu, and Ms Romina Basting.

## REFERENCES

1. Lozano R, Naghavi M, Foreman K, et al. Global and regional mortality from 235 causes of death for 20 age groups in 1990 and 2010: A systematic analysis for the Global Burden of Disease Study 2010. The Lancet. 2012;380(9859):2095–2128. doi:10.1016/S0140-6736(12)61728-0

2. Feigin VL, Forouzanfar MH, Krishnamurthi R, et al. Global and regional burden of stroke during 1990–2010: Findings from the Global Burden of Disease Study 2010. The Lancet. 2014;383(9913):245–255. doi:10.1016/S0140-6736(13)61953-4

3. Seminog OO, Scarborough P, Wright FL, Rayner M, Goldacre MJ. Determinants of the decline in mortality from acute stroke in England: Linked national database study of 795 869 adults. BMJ. 2019;365:l1778. doi:10.1136/bmj.l1778

4. Johnson CO, Nguyen M, Roth GA, et al. Global, regional, and national burden of stroke, 1990–2016: A systematic analysis for the Global Burden of Disease Study 2016. Lancet Neurol. 2019;18(5):439–458. doi:10.1016/S1474-4422(19)30034-1

5. Pendlebury ST, Rothwell PM. Incidence and prevalence of dementia associated with transient ischaemic attack and stroke: Analysis of the population-based Oxford Vascular Study. Lancet Neurol. 2019;18(3):248–258. doi:10.1016/S1474-4422(18)30442-3

6. Hurford R, Charidimou A, Fox Z, Cipolotti L, Werring DJ. Domain-specific trends in cognitive impairment after acute ischaemic stroke. J Neurol. 2013;260(1):237–241. doi:10.1007/s00415-012-6625-0

7. Lesniak M, Bak T, Czepiel W, Seniow J, Czlonkowska A. Frequency and prognostic value of cognitive disorders in stroke patients. Dement Geriatr Cogn Disord. 2008;26(4):356–363. doi:10.1159/000162262

8. Zinn S, Bosworth HB, Hoenig HM, Swartzwelder HS. Executive function deficits in acute stroke. Arch Phys Med Rehabil. 2007;88(2):173–180. doi:10.1016/j.apmr.2006.11.015

9. Pohjasvaara T, Leskelä M, Vataja R, et al. Post-stroke depression, executive dysfunction and functional outcome. Eur J Neurol. 2002;9(3):269–275. doi:10.1046/j.1468-1331.2002.00396.x

10. Mole JA, Demeyere N. The relationship between early post-stroke cognition and longer term activities and participation: A systematic review. Neuropsychol Rehabil. 2020;30(2):346–370. doi:10.1080/09602011.2018.1464934

11. Melkas S, Vataja R, Oksala NKJ, et al. Depression–executive dysfunction syndrome relates to poor poststroke survival. Am J Geriatr Psychiatry. 2010;18(11):1007–1016. doi:10.1097/JGP.0b013e3181d695d7

12. Muir RT, Lam B, Honjo K, et al. Trail Making Test elucidates neural substrates of specific poststroke executive dysfunctions. Stroke. 2015;46(10):2755–2761. doi:10.1161/STROKEAHA.115.009936

13. Veldsman M, Werden E, Egorova N, Khlif MS, Brodtmann A. Microstructural degeneration and cerebrovascular risk burden underlying executive dysfunction after stroke. Sci Rep. 2020;10(1):17911. doi:10.1038/s41598-020-75074-w

14. Ihle-Hansen H, Thommessen B, Fagerland M, et al. Impact of white matter lesions on cognition in stroke patients free from pre-stroke cognitive impairment: A one-year follow-up study. Dement Geriatr Cogn Disord Extra. 2012;2:38–47. doi:10.1159/000336817

15. Fu JH. Extent of white matter lesions is related to acute subcortical infarcts and predicts further stroke risk in patients with first ever ischaemic stroke. J Neurol Neurosurg Psychiatry. 2005;76(6):793–796. doi:10.1136/jnnp.2003.032771

16. Li L, Simoni M, Küker W, et al. Population-based case-control study of white matter changes on brain imaging in transient ischemic attack and ischemic stroke. Stroke. 2013;44(11):3063–3070. doi:10.1161/STROKEAHA.113.002775

17. Singer OC, Sitzer M, du Mesnil de Rochemont R, Neumann-Haefelin T. Practical limitations of acute stroke MRI due to patient-related problems. Neurology. 2004;62(10):1848. doi:10.1212/01.WNL.0000125320.53244.FA

18. Demeyere N, Riddoch MJ, Slavkova ED, Bickerton WL, Humphreys GW. The Oxford Cognitive Screen (OCS): Validation of a stroke-specific short cognitive screening tool. Psychol Assess. 2015;27(3):883–894. doi:10.1037/pas0000082

19. Demeyere N, Haupt M, Webb SS, et al. Introducing the tablet-based Oxford Cognitive Screen-Plus (OCS-Plus) as an assessment tool for subtle cognitive impairments. Sci Rep. 2021;11(1):8000. doi:10.1038/s41598-021-87287-8

20. Humphreys GW, Duta MD, Montana L, et al. Cognitive function in low-income and low-literacy settings: Validation of the tablet-based Oxford Cognitive Screen in the health and aging in Africa: A longitudinal study of an INDEPTH Community in South Africa (HAALSI). J Gerontol Ser B. 2017;72(1):38–50. doi:10.1093/geronb/gbw139

21. Burgess PW, Shallice T. Bizarre responses, rule detection and frontal lobe lesions. Cortex. 1996;32(2):241–259. doi:10.1016/S0010-9452(96)80049-9

22. Moore M. A practical guide to lesion symptom mapping. Published online 2022. doi:10.31234/osf.io/2jxr9

23. Rorden C, Bonilha L, Fridriksson J, Bender B, Karnath HO. Age-specific CT and MRI templates for spatial normalization. NeuroImage. 2012;61(4):957–965. doi:10.1016/j.neuroimage.2012.03.020

24. Griffis JC, Metcalf NV, Corbetta M, Shulman GL. Lesion Quantification Toolkit: A MATLAB software tool for estimating grey matter damage and white matter disconnections in patients with focal brain lesions. NeuroImage Clin. 2021;30:102639. doi:10.1016/j.nicl.2021.102639

25. Yeh FC, Panesar S, Fernandes D, et al. Population-averaged atlas of the macroscale human structural connectome and its network topology. NeuroImage. 2018;178:57–68. doi:10.1016/j.neuroimage.2018.05.027

26. Wahlund LO, Barkhof F, Fazekas F, et al. A new rating scale for age-related white matter changes applicable to MRI and CT. Stroke. 2001;32(6):1318–1322. doi:10.1161/01.STR.32.6.1318

27. Simoni M, Li L, Paul NLM, et al. Age- and sex-specific rates of leukoaraiosis in TIA and stroke patients. Neurology. 2012;79(12):1215–1222. doi:10.1212/WNL.0b013e31826b951e

28. Cho MJ, Jang SH. Relationship between post-traumatic amnesia and white matter integrity in traumatic brain injury using tract-based spatial statistics. Sci Rep. 2021;11(1):6898. doi:10.1038/s41598-021-86439-0

29. Kamali A, Kramer LA, Butler IJ, Hasan KM. Diffusion tensor tractography of the somatosensory system in the human brainstem: Initial findings using high isotropic spatial resolution at 3.0 T. Eur Radiol. 2009;19(6):1480–1488. doi:10.1007/s00330-009-1305-x

30. Santiago C, Herrmann N, Swardfager W, et al. White matter microstructural integrity is associated with executive function and processing speed in older adults with coronary artery disease. Am J Geriatr Psychiatry. 2015;23(7):754–763. doi:10.1016/j.jagp.2014.09.008

31. Kantarci K, Senjem ML, Avula R, et al. Diffusion tensor imaging and cognitive function in older adults with no dementia. Neurology. 2011;77(1):26–34. doi:10.1212/WNL.0b013e31822313dc

32. Charlton RA, Schiavone F, Barrick TR, Morris RG, Markus HS. Diffusion tensor imaging detects age related white matter change over a 2 year follow-up which is associated with working memory decline. J Neurol Neurosurg Psychiatry. 2010;81(1):13–19. doi:10.1136/jnnp.2008.167288

33. Bettcher BM, Mungas D, Patel N, et al. Neuroanatomical substrates of executive functions: Beyond prefrontal structures. Neuropsychologia. 2016;85:100–109. doi:10.1016/j.neuropsychologia.2016.03.001

34. Cristofori I, Zhong W, Chau A, Solomon J, Krueger F, Grafman J. White and gray matter contributions to executive function recovery after traumatic brain injury. Neurology. 2015;84(14):1394–1401. doi:10.1212/WNL.0000000000001446

35. Pendlebury ST, Rothwell PM. Prevalence, incidence, and factors associated with pre-stroke and post-stroke dementia: A systematic review and meta-analysis. Lancet Neurol. 2009;8(11):1006–1018. doi:10.1016/S1474-4422(09)70236-4

36. Wippold FJ. Head and neck imaging: The role of CT and MRI. J Magn Reson Imaging. 2007;25(3):453–465. doi:10.1002/jmri.20838

37. Pendlebury ST, Thomson RJ, Welch SJV, Kuker W, Rothwell PM, for the Oxford Vascular Study. Utility of white matter disease and atrophy on routinely acquired brain imaging for prediction of long-term delirium risk: Population-based cohort study. Age Ageing. 2022;51(1):afab200. doi:10.1093/ageing/afab200

38. Ferguson KJ, Cvoro V, MacLullich AMJ, et al. Visual rating scales of white matter hyperintensities and atrophy: Comparison of computed tomography and magnetic resonance imaging. J Stroke Cerebrovasc Dis. 2018;27(7):1815–1821. doi:10.1016/j.jstrokecerebrovasdis.2018.02.028

39. Dostovic Z, Dostovic E, Smajlovic D, Ibrahimagic OC, Avdic L. Brain edema after ischaemic stroke. Med Arch. 2016;70(5):339–341. doi:10.5455/medarh.2016.70.339-341

40. Griffanti L, Zamboni G, Khan A, et al. BIANCA (Brain Intensity AbNormality Classification Algorithm): A new tool for automated segmentation of white matter hyperintensities. NeuroImage. 2016;141:191–205. doi:10.1016/j.neuroimage.2016.07.018

