## Supplementary Materials for "Post-stroke executive function in relation to white matter damage on clinically acquired CT brain imaging"

**Supplementary Figure 1.** OCS-Plus Rule Finding task. Patients are shown three columns of alternating geometric shapes (squares-triangles-squares), rows of alternating luminosity (dark–light), and a red dot that moves around the pattern following certain rules. Patients must try to learn the rules to predict where the dot is going to go next based on previous moves. Patients must tap the shape with a stylus to respond.

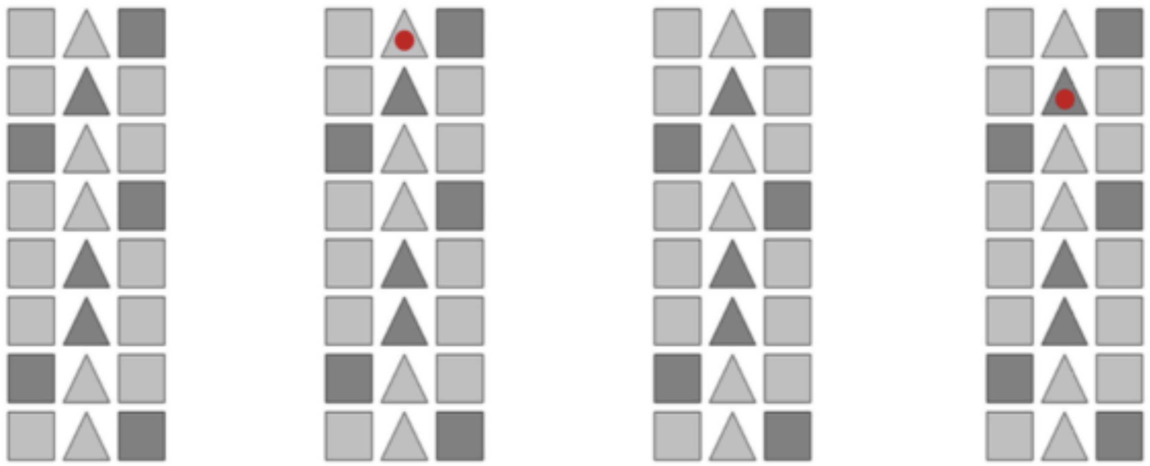

**Supplementary Table 1.** Visual rating criteria for the Age-Related White Matter Changes scale. WMHs on CT were defined as hypodense areas of ≥5 mm.

| | **0** | **1** | **2** | **3** |
|---|---|---|---|---|
| **Frontal**<br>**Parieto-occipital**<br>**Temporal**<br>**Infratentorial** | No lesions (including symmetrical, well-defined caps or bands). | Focal lesions. | Beginning confluence of lesions. | Diffuse involvement of the entire region, with or without U-fibres. |
| **Basal ganglia** | No lesions. | Single focal lesion. | Two or more focal lesions. | Confluent lesions. |

**Supplementary Table 2.** Summary model statistics from the multiple linear regression analyses conducted in the present study to investigate the association between post-stroke executive function and (1) overall stroke damage to the white matter, (2) stroke damage to individual white matter tracts, (3) white matter damage associated with WMHs, (4) white matter damage associated with WMHs and stroke damage to medial lemniscus.

| Model | *F* | *Multiple* $R^2$ | *Adjusted* $R^2$ | *p* |
|---|---|---|---|---|
| **1) Overall stroke damage to white matter** | 2.56 | 0.37 | 0.22 | .004* |
| **2) Stroke damage to individual white matter tracts** | | | | |
| Anterior commissure | 2.61 | 0.37 | 0.23 | .003* |
| Arcuate fasciculus | 2.38 | 0.35 | 0.20 | .007* |
| Acoustic radiation | 2.57 | 0.37 | 0.23 | .003* |
| Cerebellum | 2.60 | 0.37 | 0.23 | .003* |
| Corpus callosum (anterior) | 2.43 | 0.36 | 0.21 | .006* |
| Corpus callosum (mid-anterior) | 2.38 | 0.35 | 0.20 | .007* |
| Corpus callosum (central) | 2.41 | 0.35 | 0.21 | .006* |
| Corpus callosum (mid-posterior) | 2.38 | 0.35 | 0.20 | .007* |
| Corpus callosum (posterior) | 2.78 | 0.39 | 0.25 | .002* |
| Corticospinal tract | 2.40 | 0.35 | 0.21 | .006* |
| Corticostriatal tract | 2.37 | 0.35 | 0.20 | .007* |
| Central tegmental tract | 2.38 | 0.35 | 0.20 | .007* |
| Corticothalamic pathway | 2.72 | 0.38 | 0.24 | .002* |
| Cingulum | 2.50 | 0.36 | 0.22 | .004* |
| Dorsal longitudinal fasciculus | 2.39 | 0.35 | 0.21 | .006* |
| Extreme capsule | 2.42 | 0.36 | 0.21 | .006* |
| Frontal aslant tract | 2.42 | 0.36 | 0.21 | .006* |
| Frontopontine tract | 2.38 | 0.35 | 0.20 | .007* |
| Fornix | 2.75 | 0.39 | 0.25 | .002* |
| Inferior cerebellar penduncle | 2.56 | 0.37 | 0.22 | .004* |
| Inferior fronto-occipital fasciculus | 2.59 | 0.37 | 0.23 | .003* |
| Inferior longitudinal fasciculus | 2.80 | 0.39 | 0.25 | .002* |
| Lateral lemniscus | 2.64 | 0.38 | 0.23 | .003* |
| Middle cerebellar peduncle | 2.84 | 0.39 | 0.26 | .001* |
| Medial longitudinal fasciculus | 2.43 | 0.36 | 0.21 | .006* |
| Medial lemniscus | 3.47 | 0.44 | 0.31 | <.001** |
| Middle longitudinal fasciculus | 2.38 | 0.35 | 0.20 | .007* |
| Occipitopontine tract | 2.95 | 0.40 | 0.27 | <.001** |
| Optic radiation | 2.53 | 0.37 | 0.22 | .004* |
| Posterior commissure | 2.42 | 0.36 | 0.21 | .005* |
| Parietopontine tract | 2.76 | 0.39 | 0.25 | .002* |
| Reticulospinal tract | 2.40 | 0.35 | 0.21 | .006* |
| Superior cerebellar peduncle | 2.38 | 0.35 | 0.20 | .007* |
| Superior longitudinal fasciculus | 2.39 | 0.35 | 0.20 | .007* |
| Spinothalamic tract | 3.22 | 0.42 | 0.29 | <.001** |
| Temporopontine tract | 3.15 | 0.42 | 0.29 | <.001** |
| Uncinate fasciculus | 2.40 | 0.35 | 0.21 | .006* |
| U fibres | 2.53 | 0.37 | 0.22 | .004* |
| Vermis | 2.88 | 0.40 | 0.26 | .001* |
| Vertical occipital fasciculus | 2.43 | 0.36 | 0.21 | .006* |
| **3) White matter damage associated with WMHs** | 2.79 | 0.39 | 0.25 | .002* |
| **4) White matter damage associated with WMHs and medial lemniscus** | 3.86 | 0.51 | 0.37 | <.001** |

*Note.* Age was a significant negative predictor of executive functioning in all models at $p < .001$. Stroke volume was a significant negative predictor of executive functioning in all models at $p < .05$. * significant at $p < .05$. ** significant at $p < .001$.
